# Development of a questionnaire library for rapid health data acquisition during wildfire events

**DOI:** 10.1101/2025.02.10.25321891

**Authors:** Lilian Liu, Christine Loftus, Diana Rohlman, Edmund Seto, Elena Austin

**Affiliations:** Department of Environmental and Occupational Health Sciences, University of Washington, Seattle, WA 98105, United States; Department of Environmental and Molecular Toxicology, Oregon State University, Corvallis, OR 97331, United States

**Keywords:** Wildfire, smoke, perishable data, health assessment, data library, questionnaire, survey, instrumentation, disaster response research, data collection

## Abstract

Rapid health outcome data acquisition using existing questionnaires can greatly facilitate time-sensitive research during or after a wildfire event. We aimed to develop a readily available library of questionnaires for self-reported health outcomes to serve as a centralized platform for wildfire researchers seeking to quickly design instruments tailored to their research aims. In this paper, we describe the methodology used to identify relevant health questionnaires and compile them into a structured library. We first followed the Preferred Reporting Items for Systematic reviews and Meta-Analyses (PRISMA) 2020 checklist and performed a systematic literature review of prior research on wildfire exposure and health and used this to 1) identify health outcome categories associated with wildfire and smoke exposure and 2) extract questionnaires used for self-report of health outcomes related to wildfire exposure. We also performed a secondary search of existing questionnaire repositories to identify additional instruments relevant to health impacts of wildfire exposure. All questionnaires (n=100) were organized by eight health outcome categories (mental health = 60, respiratory health = 19, overall health = 17, sleep = 10, cardiovascular health = 4, allergy = 1, irritation (eye, throat, skin) = 2, and metabolic health = 1). The [library](https://www.medrxiv.org/content/medrxiv/early/2025/02/13/2025.02.10.25321891/DC1/embed/media-1.xlsx?download=true) (see supplementary information) will be accompanied by a decision-tree framework in development, which will assist future users in building new questionnaires that best fit their study population and research aims. Both the questionnaire library and the forthcoming decision-support framework will be publicly accessible to researchers, public health agencies, and other groups interested in rapid response data collection to characterize the impacts of wildfires. Additionally, this method of creating a wildfire exposure health questionnaire library may serve as a template for rapid collection of questionnaire-based data following other disasters.

## Background

### 1.1 Wildfire events, and the need for rapid collection of perishable questionnaire-based data

Wildfire (WF) events have increased in frequency, intensity, and duration in recent years in the United States (Dennison et al., 2014; Salguero et al., 2020). Exposure to both fires and WF-related smoke contributes to various health outcomes (Finlay et al., 2012; Rizzo & Rizzo, 2024) and leads to numerous public health concerns (Reisen et al., 2015). To characterize WF-related health impacts in defined population, researchers usually acquire health data using various methods, including use of administrative records (e.g., hospitalization records, insurance claims, or pharmaceutical records), surveillance/census data, biospecimen collection and analyses, clinical exams and assessments, crowdsourced data (e.g. secondary data from commercial companies (e.g. Lumos Lab) or research frameworks (e.g. Apple ResearchKit)), and administration of questionnaires for self-reported impacts (Barkoski et al., 2024). Due to the rapidly evolving nature of disasters, such as of WF events, health data are often perishable (Adams et al., 2024) and need to be collected in a timely fashion post-disaster. Questionnaires are useful tools for ascertainment of self-reported health impacts, but *de novo* questionnaire design can be slow, impeding rapid deployment of surveys and increasing the chances of limited accuracy due to recall. Moreover, in times of disasters, there is a need to identify the minimum relevant set of questions needed for research to not overburden WF affected study populations. Additionally, data collected by different research groups may not always be comparable to each other as the data collection processes vary. This would likely lead to less scientific discovery and interpretation of results due to inconsistent data collection.

To address the need for WF-related questionnaires and to encourage reproducible data collection research design, we are developing a decision-support framework to help WF researchers identify a library of relevant questionnaires specific to the WF context and the research goals. This decision-support framework aims to facilitate rapid and reproducible questionnaire design for perishable WF-related human exposure and health outcome data acquisition. Here we present an intermediate product of the framework –a centralized platform consisting of WF-relevant health questionnaires gathered from a literature review and search of existing libraries of health questionnaires – that can be used either with the decision-support framework or as a standalone resource to support time-sensitive WF research.

### 1.2 Objectives and aims

Rapid data acquisition is essential to support effective disaster research, such as research related to the health impacts of WF events (Gorbea Díaz et al., 2023; Morton & Levy, 2011). Therefore, having a centralized health questionnaire library built within a decision-support framework can help support time-sensitive WF research. This methods article describes the process of acquiring and creating a library of health questionnaires, which is a core component of our forthcoming decision-support framework for WF research. Our methods, described herein, are intended to serve as methodological documentation to support the released of the framework and its health questionnaire library as a public resource.

## Method details

### 2.1 Literature review and health outcome determination

In this study, we define WF exposure as impacts of WFs in a nonoccupational context, including fires due to non-prescribed burns or wildland-urban interface (WUI) fire, as well as exposure to WF smoke from either a local WF event or migrated smoke from a WF event afar. We will “WF” hereafter to represent both WF and WF smoke. In addition to the generic health impacts, because increased asthma symptoms are an important health outcome associated with WF exposure (Balmes et al., 2024; Noah et al., 2023), we decided to solicit studies that reported respiratory health impact from WF exposure including asthma. To identify relevant published literature, we used a combination of search terms. The search term combination must include at least one word from the three topics, including 1) “wildfire” or “wildfire smoke”, 2a) “exposure” or 2b) “health”, “impact(s)/effect(s)”, or “asthma/respiratory”, and 3) “assessment”. We applied the search term combinations (n = 14) on Web of Science and PubMed and exported results as .txt files to the online systematic review tool Covidence. To screen for eligible articles, we applied inclusion criteria, including that the aim of the paper was on non-occupational WF exposure/health outcomes assessment, the paper described questionnaires for collecting reported WF-related human exposure and/or health outcome data, and the paper was published in English. Exclusion criteria include no questionnaire-based assessments of human WF exposure or health outcomes conducted, only non-English questionnaires provided, focus on WF exposure in occupational context only, perceived risks without presence of WF (e.g. likelihood of a neighborhood of future WF event), property damage, land management, air treatment efficacy, or exposure reduction/ health therapeutic effects (e.g. air treatment technology efficacy). We included questionnaires regardless of administration (e.g., self-administered vs. administered by research team staff). We found 878 studies after removing 3,049 duplicates. After screening based on title and abstract, 106 were assessed for full-text screening and ultimately, 30 studies (Table S1) (Adu MK et al., 2022, Adu MK et al., 2024; Agyapong B, Shalaby R, Eboreime E, Obuobi-Donkor G, et al., 2022; Agyapong B, Shalaby R, Eboreime E, Wei Y, et al., 2022; Agyapong et al., 2022; Agyapong VIO et al., 2020; Brown et al., 2019; Brown MRG et al., 2021; Bryant RA et al., 2018; Castillo C et al., 2024; Cherry N & Haynes W, 2017; Cowlishaw S et al., 2021; Haikerwal A et al., 2021; Hoshiko S et al., 2023; Künzli et al., 2006; Mao W et al., 2022, Mao W et al., 2024; Marshall et al., 2007; Mirabelli et al., 2009; Mirabelli MC et al., 2022; Moosavi S et al., 2019; Obuobi-Donkor G et al., 2024; Pacella BJ et al., 2024; Papadatou et al., 2012; Papanikolaou V et al., 2011; Psarros C et al., 2017; Ritchie A et al., 2021; Rodney RM et al., 2021; Usher K et al., 2022; Wasiak J et al., 2013) met all inclusion criteria and were selected for data extraction. A Preferred Reporting Items for Systematic reviews and Meta-Analyses (PRISMA) flow diagram (Fig. 1) provides details about the literature review process, which was conducted in Covidence. Additionally, we separately searched for asthma-related questionnaires in the Research Electronic Data Capture (REDCap) Shared Library for the same reason for literature review searches.

**Figure 1.**
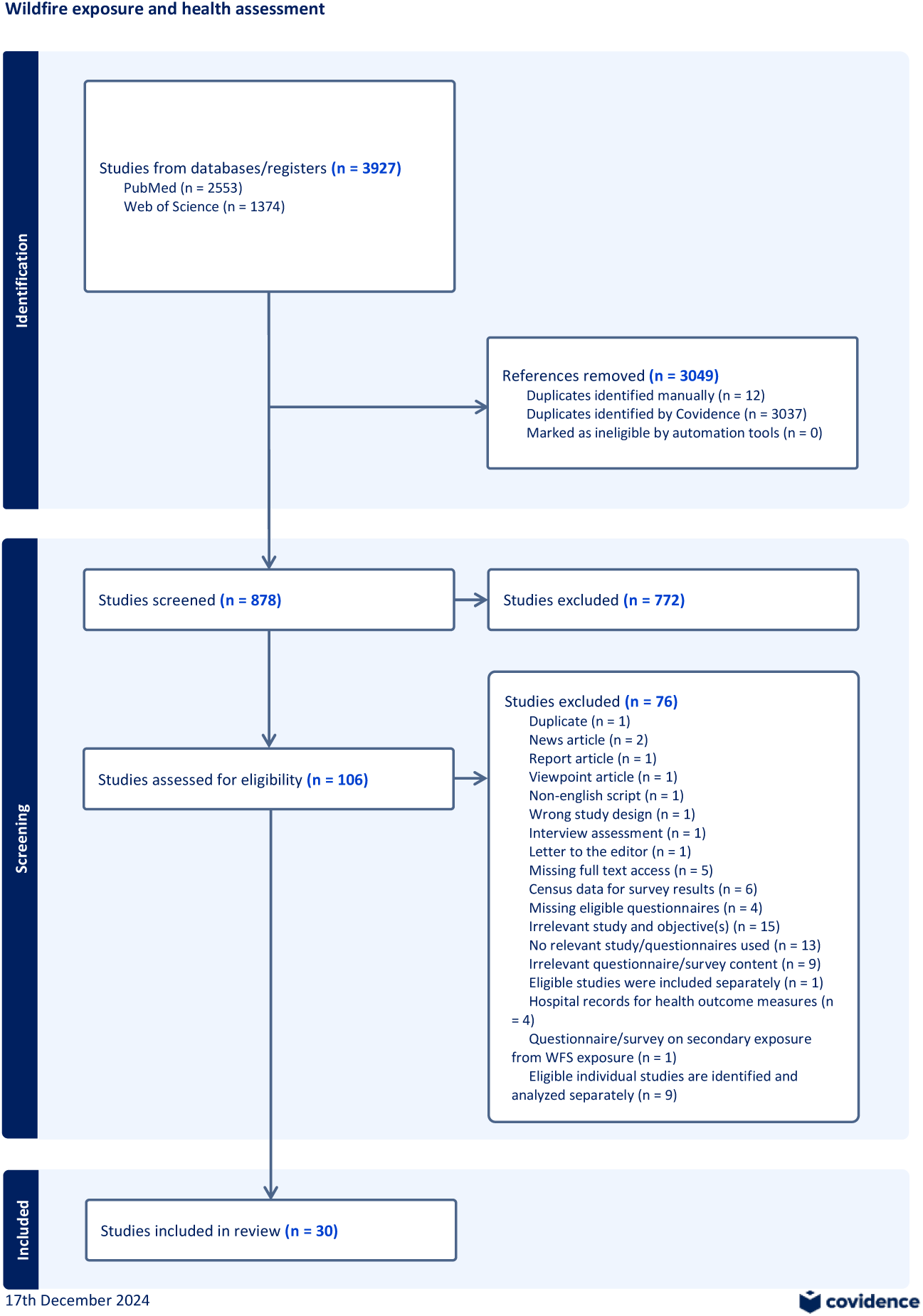
PRISMA flow diagram of literature review results.

During the literature review data extraction step, we extracted all full-text questionnaires used to collect information on reported health impacts during or after WF events and categorized them based on the health outcome(s) assessed. Based on our eligibility criteria, included questionnaires/questionnaire sections (i.e. for questionnaires with multiple sections addressing different health outcomes, we considered that section as its own questionnaire to facilitate organization by health outcome category) assess health outcomes that are from direct impact of WF exposure. We will use “questionnaire(s)” hereafter to represent questionnaires/questionnaire sections. For example, a questionnaire may ask questions like “during or immediately after the fire, did you experience any health problems caused or made worse by the fires?” (Cherry N & Haynes W, 2017). Additionally, we separately searched for asthma-related questionnaires in the Research Electronic Data Capture (REDCap) Shared Library for the same reason for literature review searches. Overall, we identified nine health outcome categories: mental health, respiratory health, sleep, cardiovascular health, allergies, irritation (eye, throat, skin), metabolic health (e.g. type II diabetes), and overall health. A total of 42 questionnaires that cover these health outcome categories were found in the research articles, and 14 asthma questionnaires were found in the REDCap Shared Library.

### 2.2. Extended search for health questionnaires

Because our literature review approach could miss many questionnaires that are commonly used in public health research outside the literature specific to WF impacts, we additionally searched for relevant health questionnaires on REDCap Shared Library as REDCap performs questionnaire acquisition twice annually from other online repositories (e.g. PROMIS). Upon review, the decision was made to directly search for other existing questionnaires related to the health outcomes of interest. These questionnaires were considered to be of high value to WF researchers. Therefore, we used the health outcome categories identified in 2.1 as keywords (Table 2) and searched existing health questionnaires on REDCap Shared Library. We screened all search results and included only questionnaires that are useful for WF research. For example, we would include a questionnaire if it asked “in the past 7 days, my sleep was restful.” with Likert ranking options because the question specifically relates to the topic of sleep. Questionnaires that did not contain questions relevant to the health outcomes (Table 1), or that relied solely on clinical measurements and diagnoses (e.g. “severity of asthma exacerbation expressed in forced expiratory volume in one second (FEV1) %”) were excluded.

**Table 1.**
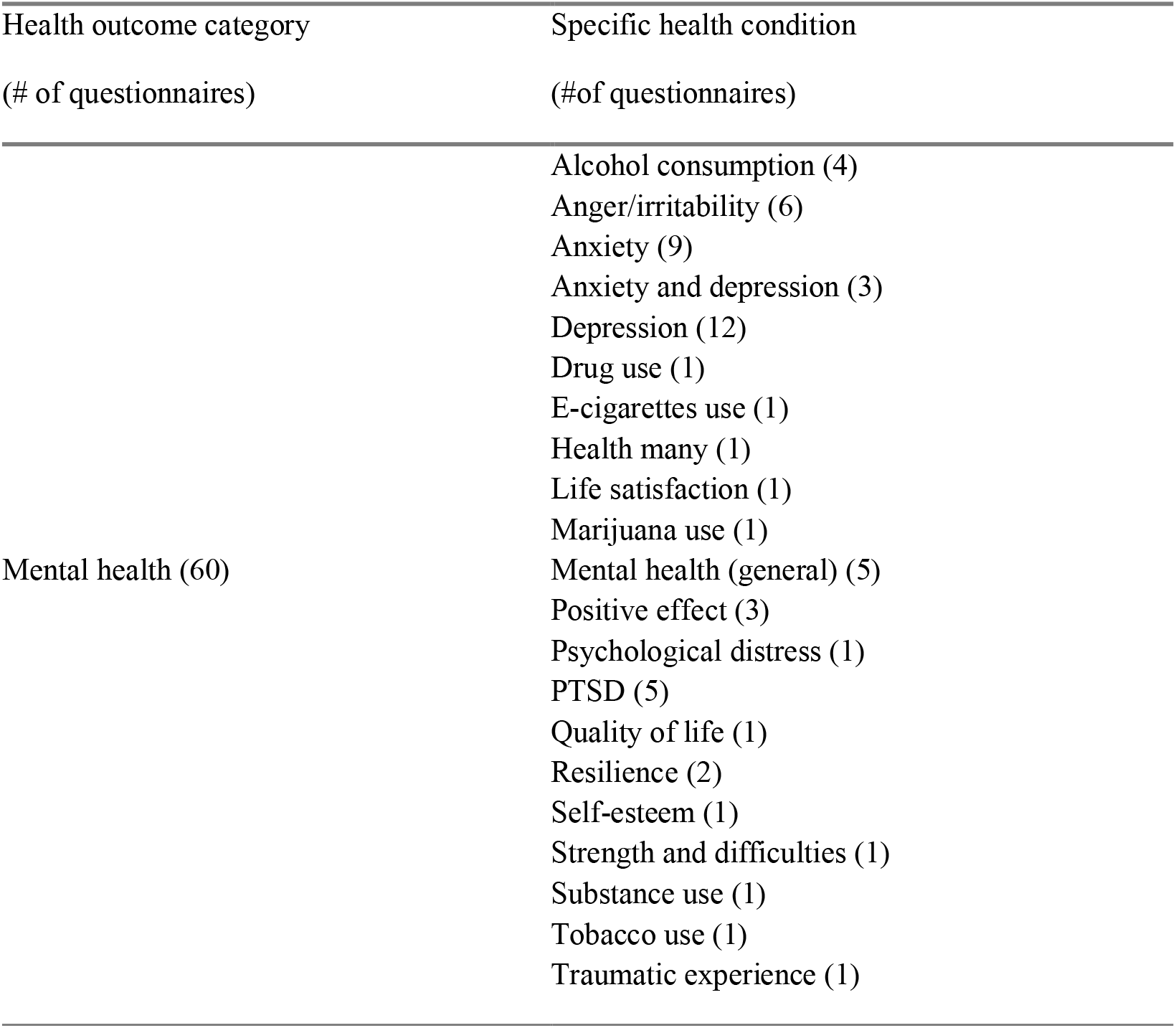
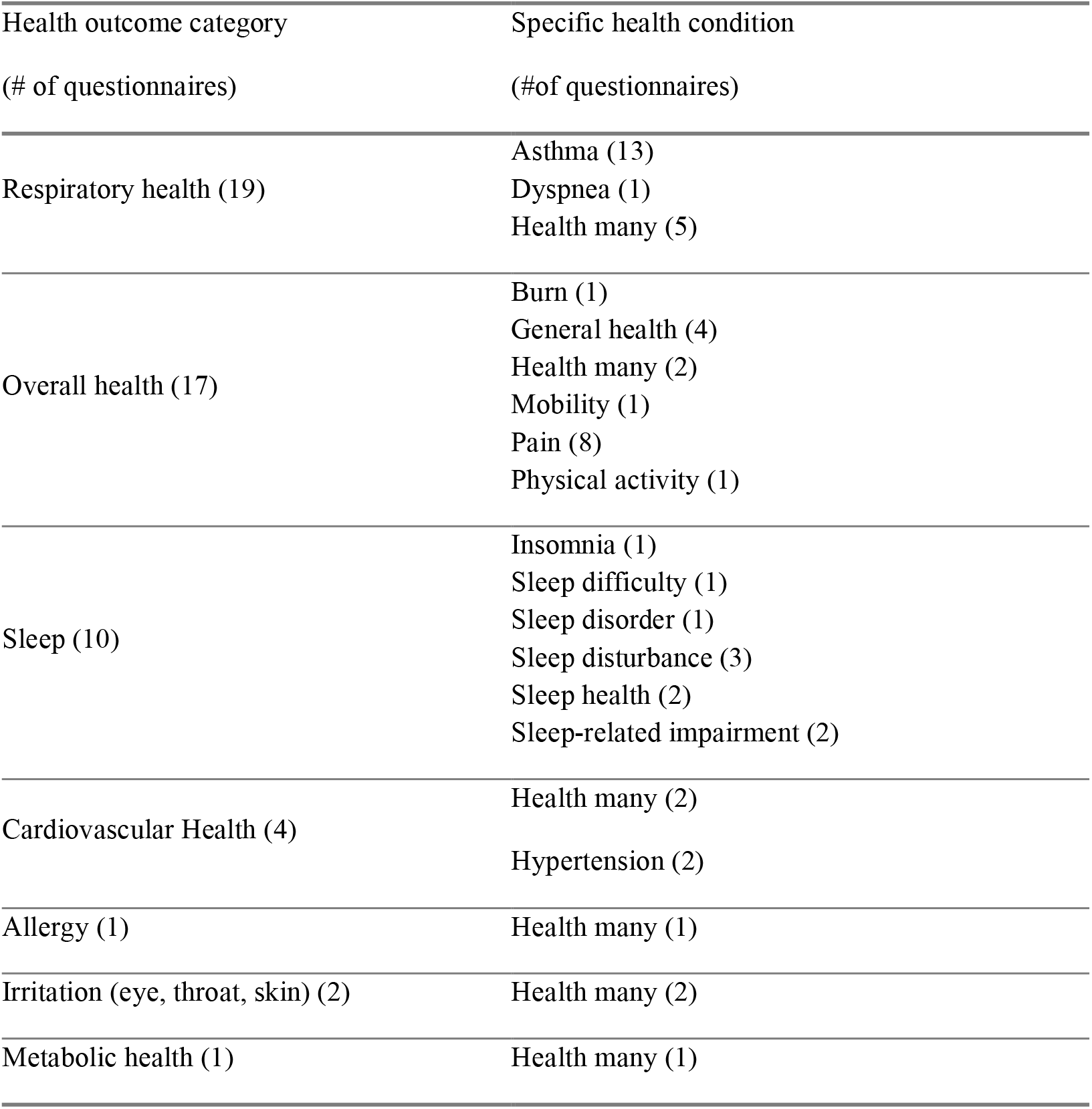
Summary of questionnaire/questionnaire sections on different health outcome assessments.

**Table 2.**
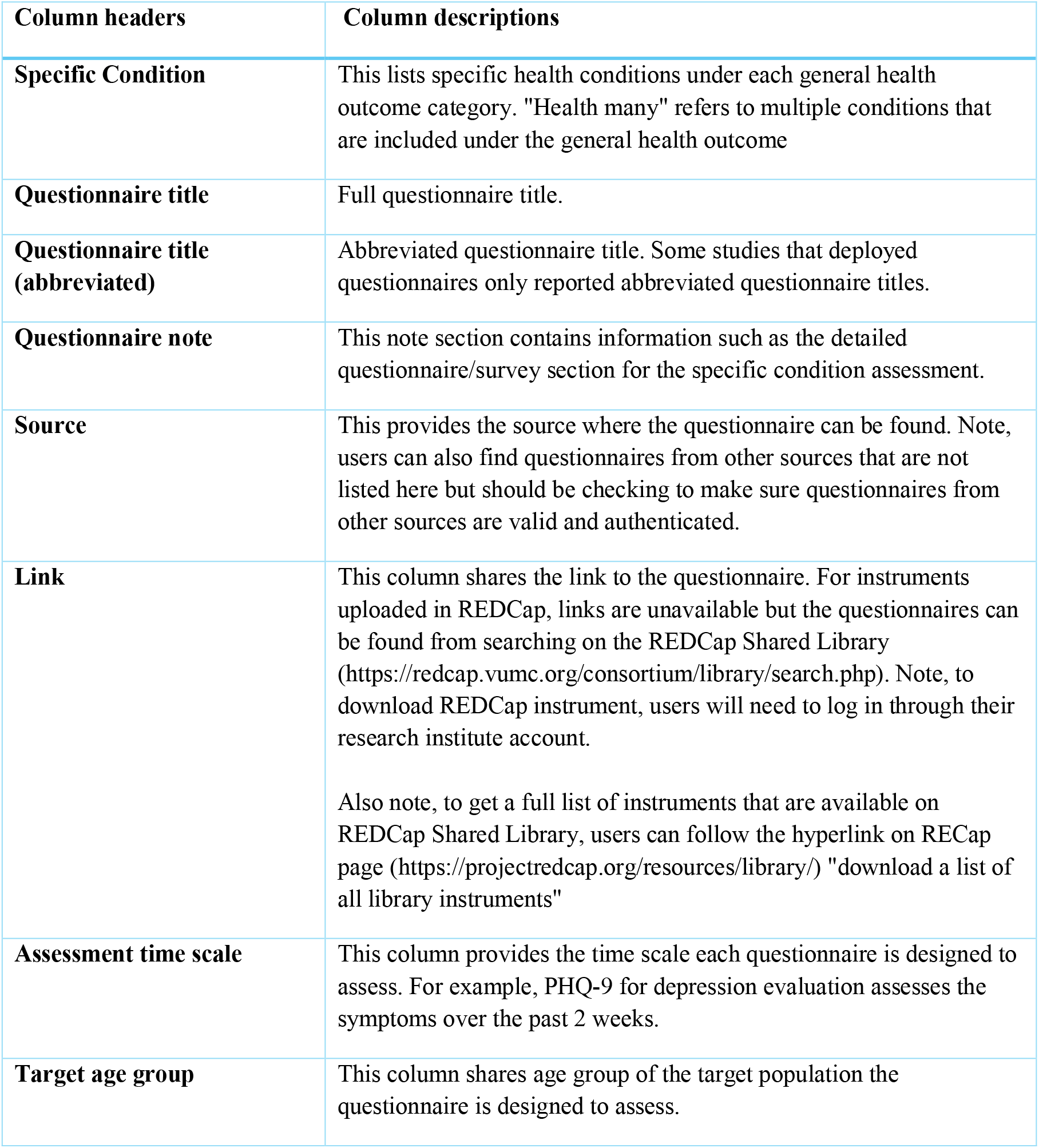
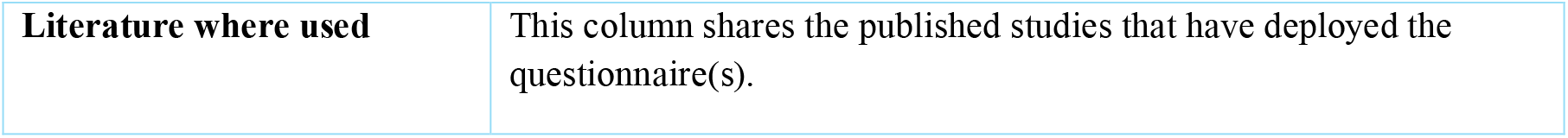
Health questionnaire library column headers and descriptions.

We found an additional 58 questionnaires assessing specific health conditions from the health outcome categories and finalized the library with a total of 100 questionnaires (Table 1). Importantly, this secondary search identified questionnaires that evaluate health effects in adolescent and younger children, enabling assessment of WF exposure and health effects across the lifespan.

### 2.3 Health questionnaire library

We organized all questionnaire content into an Excel spreadsheet with the first worksheet providing general information and user instructions and the following worksheets listing questionnaires by health outcome categories. For each questionnaire, we provided: specific health condition assessed, titles (full and abbreviated), questionnaire note, source, website link (if applicable), assessment reference period (e.g., last 7 days, etc.), target age group, and literature citing use of the questionnaire based on our literature review (Table 2). These column headers were created to help users navigate and filter the options. These worksheets will be updated periodically by acquiring and integrating new questionnaires to ensure the library remains current.

The library is intended to be made available as an interactive online resource that is openly accessible. The library can be used as a standalone tool, or in combination with the decision-support framework. By going directly to the library, users can view and filter the questionnaires based on their unique research needs, and acquire them by visiting the link provided, viewing the cited papers, or by searching for the questionnaire title in a web search engine. The added benefit of having the library is to facilitate WF research design, as the content in the library may help researchers brainstorm health outcomes that would potentially apply to their study objectives and enabling further scientific discovery by serving as a standard tool that allow consistent data collection design for later data sharing and data reuse. Additionally, the reuse of existing questionnaires from other WF studies may promote consistency between studies, allowing for comparisons of findings between WF events, which would enhance overall reliability and reproducibility of WF research.

## Method validation

### 3.1 Health outcomes

As part of a decision-support framework, this library of questionnaires provides WF researchers with a centralized platform to browse and acquire relevant instruments for their studies and is available as a spreadsheet (Supplemental Materials). The creation of this library is in direct response to researcher needs; a survey of researchers within the National Institute of Environmental Sciences (NIEHS) Disaster Response Research (DR2) network revealed a need for tools to enable rapid data acquisition, and a library of health questionnaires. Our library addresses both needs; users of the library can browse and acquire a variety of health questionnaires to expedite their research by deploying readily available instruments.

The library has been made available to the Public Health Extreme Events Research (PHEER) group, which is a *“public health researcher-led network that can mobilize rapidly, inform evolving disaster research agendas and funding decisions, and advance the field of public health disaster science*”. Currently, PHEER is responding to the January 2025 wildfire events in Los Angeles, California, USA. Our library is supporting time-sensitive mobilization efforts by providing existing questionnaire instruments to facilitate perishable WF health outcome data acquisition.

### Limitations

We conducted a robust literature review, supplemented by a search of health specific questionnaire repositories to identify questionnaires that could be used to assess impacts of WF exposure in humans. However, we acknowledge several limitations with our approach:

1. Validity of questionnaires: Not all included questionnaires have undergone formal validation. However, these questionnaires have been deployed in wildfire research settings and used to collect perishable data. During our periodic health questionnaire library maintenance, we will search for and add validation status to questionnaires and continue to expand the library over time. Researchers are advised to verify the suitability of each questionnaire before deployment.
2. Limited scope of search in identifying questionnaires: The questionnaire library was compiled with health questionnaires identified from a structure literature review and extended research using what was found as most relevant health outcome categories from the literature review result. Factors such as database restriction, language limitations, exclusion of unpublished instruments, limited searches in single online questionnaire repository (i.e. REDCap Shared Library) may have contributed to gaps in coverage of the literature review results and health questionnaire acquisition. Future maintenance of the library should expand search strategies to include additional sources such as systematic reviews, government agencies, and international datasets, as well as expand the health questionnaire acquisition using various online repositories other than REDCap Shared Library.
3. Limited guidance on questionnaire adaptations: While the library includes a range of questionnaires covering multiple health outcomes, there is no explicit guidance on how researchers can adapt these instruments to different populations (e.g., pediatric groups, non-English speakers). Researchers should consider the cultural and contextual appropriateness of each instrument before application Future updates should incorporate recommendations on questionnaire modifications while maintaining psychometric integrity.
4. Lack of structured evaluation: The usability and effectiveness of the library has not yet been formally assessed. While the library has been curated based on expert discussion and review, no structured evaluation has been conducted to assess alignment with WF researchers’ needs in real time. Future work should include pilot and feasibility testing with WF researchers to refine the library based on practical usability feedback.

Given the ongoing large scale WF events, such as the 2025 Los Angeles wildfires, repositories such as ours are essential for supporting rapid human health data acquisition. Future updates will prioritize validation studies to ensure data reliability, while the literature review will be broadened to include additional relevant questionnaires. Adaptation guidelines will be developed for diverse populations, and pilot testing with wildfire researchers will enhance usability. These efforts will ensure this library remains a reliable, adaptable, and valuable tool in wildfire disaster research.

## Supporting information

Wildfire health questionnaire library SI

## Data Availability

All data produced in the present work are contained in the manuscript and supplementary information.

## Ethics statements

Not applicable.

## CRediT author statement

**Lilian Liu**: Conceptualization, Methodology, Formal analysis, Investigation, Data curation, Writing - Original draft. **Christine Loftus**: Conceptualization, Validation, Writing - Review & Editing. **Diana Rohlman**: Conceptualization, Validation, Writing - Review & Editing. **Edmund Seto**: Conceptualization, Validation, Writing - Review & Editing, Supervision. **Elena Austin**: Conceptualization, Methodology, Validation, Writing - Review & Editing, Supervision, Investigation Funding acquisition.

## Acknowledgments

Supports for this research were provided by grants from the National Institute of Environmental Health Sciences, National Institutes of Health (UW EDGE Center, P30ES007033; P30ES030287; P42ES016465). The content is solely the responsibility of the authors and does not necessarily represent the official views of the National Institutes of Health.

